# Identifying Psychosis Episodes in Psychiatric Admission Notes via Rule-based Methods, Machine Learning, and Pre-Trained Language Models

**DOI:** 10.1101/2024.03.18.24304475

**Authors:** Yining Hua, Suzanne V. Blackley, Ann K. Shinn, Joseph P. Skinner, Lauren V. Moran, Li Zhou

**Affiliations:** Departmetn of Epidemiology, T.H. Chan School of Public Health, Harvard University, Boston, Massachusetts, USA; Departmetn of Biomedical Informatics, Harvard Medical School, Boston, Massachusetts, USA; Department of General Internal Medicine, Brigham and Women’s Hospital and Harvard Medical School. Boston, Massachusetts, USA; Mass General Brigham, Somerville, Massachusetts, USA; McLean Hospital, Belmont, Massachusetts, USA; Department of Psychiatry, Harvard Medical School, Boston, Massachusetts, USA

**Keywords:** Clinical psychiatry, electronic health records, psychosis identification, machine learning, natural language processing

## Abstract

Early and accurate diagnosis is crucial for effective treatment and improved outcomes, yet identifying psychotic episodes presents significant challenges due to its complex nature and the varied presentation of symptoms among individuals. One of the primary difficulties lies in the underreporting and underdiagnosis of psychosis, compounded by the stigma surrounding mental health and the individuals’ often diminished insight into their condition. Existing efforts leveraging Electronic Health Records (EHRs) to retrospectively identify psychosis typically rely on structured data, such as medical codes and patient demographics, which frequently lack essential information. Addressing these challenges, our study leverages Natural Language Processing (NLP) algorithms to analyze psychiatric admission notes for the diagnosis of psychosis, providing a detailed evaluation of rule-based algorithms, machine learning models, and pre-trained language models. Additionally, the study investigates the effectiveness of employing keywords to streamline extensive note data before training and evaluating the models. Analyzing 4,617 initial psychiatric admission notes (1,196 cases of psychosis versus 3,433 controls) from 2005 to 2019, we discovered that the XGBoost classifier employing Term Frequency-Inverse Document Frequency (TF-IDF) features derived from notes pre-selected by expert-curated keywords, attained the highest performance with an F1 score of 0.8881 (AUROC [95% CI]: 0.9725 [0.9717, 0.9733]). BlueBERT demonstrated comparable efficacy an F1 score of 0.8841 (AUROC [95% CI]: 0.97 [0.9580, 0.9820]) on the same set of notes. Both models markedly outperformed traditional International Classification of Diseases (ICD) code-based detection methods from discharge summaries, which had an F1 score of 0.7608, thus improving the margin by 0.12. Furthermore, our findings indicate that keyword pre-selection markedly enhances the performance of both machine learning and pre-trained language models. This study illustrates the potential of NLP techniques to improve psychosis detection within admission notes and aims to serve as a foundational reference for future research on applying NLP for psychosis identification in EHR notes.

## 1. INTRODUCTION

Psychotic disorders are a category of mental disorders characterized by abnormal thoughts and perceptions and typically present with positive symptoms such as delusions and hallucinations. Accurate identification of patients experiencing psychosis is important for both clinical care and research. Considerable evidence exists of an association between duration of untreated psychosis and clinical and functional outcomes.^1,2^ For example, a 12-year prospective study of 171 patients experiencing first-episode psychosis found that at follow-up, patients who had experienced longer delays in initial treatment had, on average, poorer remission status, more severe symptoms, and greater social and quality and life impairment.^3^ However, psychosis case identification is complicated by the fact that it is often underreported and under-diagnosed.^4^ Individuals with psychosis often lack insight, leading to delays in seeking care. Moreover, on upon presentation for a first psychiatric hospitalization for recent onset psychosis, patients are often guarded and withhold key information, complicating diagnosis.

Electronic health records (EHR) offer a valuable source of information for identifying early signs of psychosis as they contain vast amounts of data on patient demographics, medical history, symptoms, and treatments that can be analyzed to identify patterns and predict outcomes.^5–7^ Although emerging research proposes to use EHRs for disease detection and prediction, most existing works related to psychosis only use structured data such as ICD-10 codes, which can be inaccurate, vague, or missing entirely.^2,3,8,9^ Meanwhile, clinical notes, among the most reliable resources for obtaining related information given their rich context, have been less studied.

However, analyzing these data using traditional statistical methods can be time-consuming and may not capture the data’s complexity and nuances. Previous research has demonstrated the potential of machine learning and deep learning techniques for accurately identifying patients with psychosis based on their structured EHR data.^10^ In a recent study conducted in South Korea, clinical data were extracted from EHRs from individuals with psychotic disorders in order to predict relapse. Three natural language processing (NLP)-enriched models were developed using three types of clinical notes (psychological tests, admission notes, and initial nursing assessment) and one complete model was developed using all three note types.^11^ Their results show that NLP models using clinical notes were more effective than models using only structured data, suggesting the importance of unstructured data in psychosis detection. In another study, NLP-derived variables of psychiatric symptoms and substance use were used to predict conversion to psychosis among patients with a prior psychiatric diagnosis.^12^ A study that combined clinical and temporal data from EHRs using a combination of rule-based information extraction and supervised machine learning methods predicted age of psychosis onset in a sample of individuals with a diagnosis of schizophrenia.^13^ However, none of these studies have utilized deep learning-based methods to analyze the EHR data despite a wealth of literature demonstrating that deep learning techniques have shown promise in clinical information studies.

One of the challenges of using deep learning techniques to analyze EHR data is the difficulty of handling the exponentially increasing computing requirements in response to the input length. To address these challenges, one way is to mitigate the noise problem by shifting the prediction level from the patient to the note section or sentence.^14^ Such approach may improve the performance of deep learning models by providing more granular context, but they also exacerbate the labeling workload, which can be labor-intensive and time-consuming.

In previous work,^15^ we validated a keyword-assisted method that uses Term Frequency-Inverse Document Frequency (TF-IDF) and machine learning classifiers to identify related keywords and extract relevant sentences before feeding them to deep learning models for the identification of patient demographics. By focusing on sentences with relevant keywords, this method minimizes the amount of irrelevant information that is fed to the deep learning models and reduces the amount of data noise. In this study, we follow the same strategy to curate psychosis-related keyword lists for noise reduction and use deep learning-based methods to address the challenges of analyzing EHR data for identifying signs of new-onset psychosis among patients hospitalized for psychiatric diagnoses. Our proposed approach has the potential to improve mental health care by identifying incident psychosis and facilitating the development of new methods for analyzing EHR data.

## 2. DATA SOURCES AND PREPARATION

### 2.1. Clinical Setting and Data Collection

This study was conducted at McLean Hospital, a psychiatric hospital in Belmont, Massachusetts, and a member of the Mass General Brigham (MGB) integrated healthcare system. All study activities were conducted with the approval of the MGB Human Research Committee (IRB) with a waiver of informed consent according to 54 U.S. Code of Federal Regulations 46.116.

The purpose of this study was to identify patients admitted to McLean for an initial psychiatric hospitalization between 2005 and 2019 and to classify patients as new onset psychosis versus another psychiatric disorder. All patients admitted to McLean are initially seen in a Clinical Evaluation Center, where they undergo a comprehensive psychiatric evaluation that is documented in the Admission Note. This centralized process ensures admission notes are similar between patients with and without psychosis. Among 21,381 patients admitted to McLean between 2005 and 2019, we excluded 10,339 patients who were older than 35 years of age, as the onset of psychosis typically occurs in adolescence and young adulthood. To identify patients with first hospitalization, we used a combination of structured data and text mining. Using structured data, we excluded patients with a previous hospitalization associated with an ICD diagnosis code for a psychiatric diagnosis within the MGB healthcare system. Figure 1 shows the process of study population identification. Future details about identification of psychosis cases are described in the next section.

**Figure 1.**
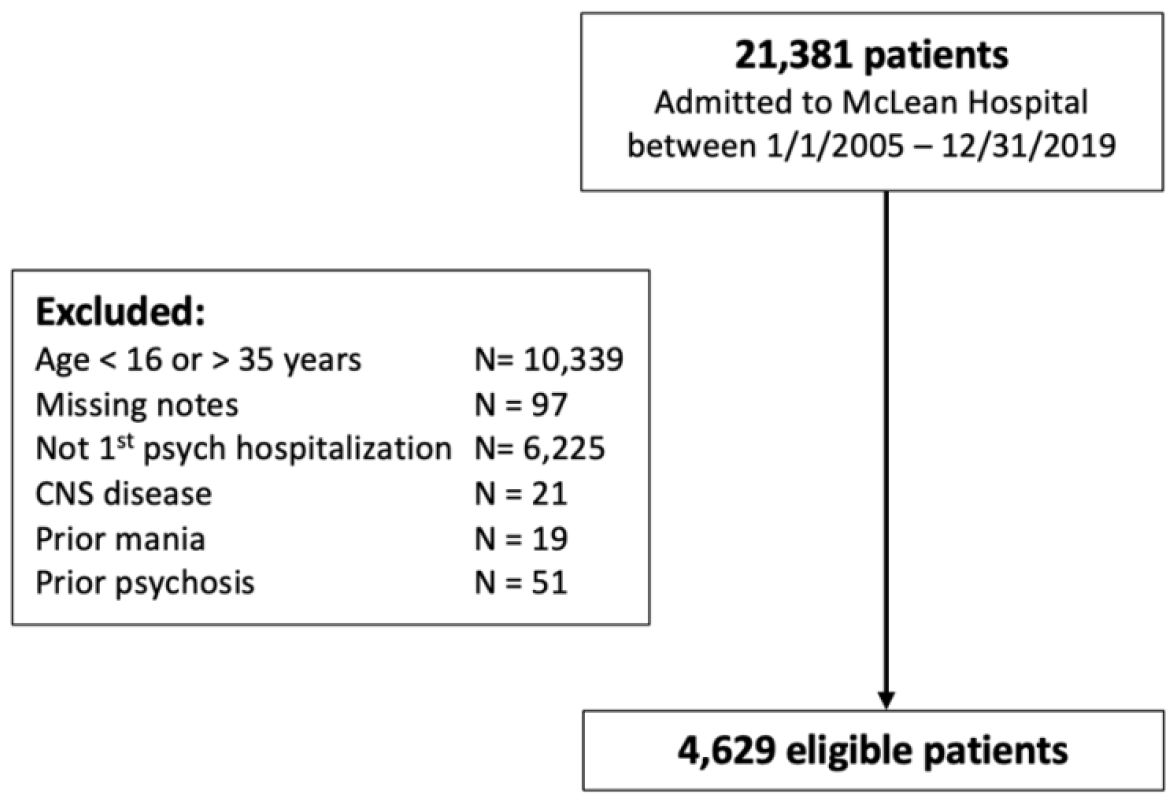
Data curation and patient cohort identification.

### 2.2. Clinician identification of psychosis

Since this healthcare system is an open system and patients may have had an initial psychiatric hospitalization in a hospital outside of MGB, we extracted admission notes. Admission notes are divided into sections that contain history of present illness that incorporates information from patient and informants (e.g., family members), past psychiatric and medical history, family history of psychiatric illness, social and developmental history, current medications and past medication trials, findings from a medical examination, and a formulation where differential diagnosis and precipitating factors are discussed, followed by a working diagnoses and initial treatment plan. We first processed admission notes by using regular expressions to de-identify sensitive information and eliminate extraneous information in the notes, including address, dates, times, provider/hospital names, patient identifiers, and zip codes, along with numerical values containing more than three digits. Addresses were masked with “[address]”, doctor names with “[doctor]”, hospital names with “[hospital]”, and dates with “[date].” Additionally, any information derived from templates and any duplicated text were systematically identified and excised from the dataset. Next, we applied text mining to identify patients with first hospitalization: Python scripts were used that searched for keywords and surrounding contexts to extract phrases for each admission note that indicated first or prior previous hospitalization and surrounding context (e.g., “multiple prior hospitalizations”, “no prior psych hospitalizations”) in past psychiatry section of note, which includes section on past hospitalizations. Phrases were manually reviewed to identify incident psychiatric hospitalizations. We excluded patients with central nervous system disease identified with ICD-9 and ICD-10 codes. We used text mining to exclude patients with past history of psychosis or mania by searching for terms and surrounding context to extract phrases consistent with new onset (e.g., “first break”) or past history of psychosis or mania (e.g., “patient with past history of schizophrenia”).

After applying the above exclusion criteria, 4,629 patients remained in the final sample. To classify patients as presenting with psychosis or another psychiatric disorder, text mining was used to identify terms related to symptoms and diagnosis of psychosis as well as words often used to describe patients with psychosis: hallucinations, delusions, psychosis/psychotic, schizo-, first break, persecutory, voice, ideas of reference, referential, thought withdrawal, broadcasting, insertion, paranoia/paranoid, loose (loosening of associations), impoverish, magic (magical thinking), flight of ideas, grandiose/grandiosity, tangential, bizarre, strange, erratic, and/or odd. The terms were selected by psychiatrists with expertise in psychosis who provided clinical care on inpatient units and were familiar with McLean admission notes. For each patient, phrases for each instance of one of the keywords and surrounding context were extracted. Phrases were manually reviewed by two psychiatrists with expertise in psychosis (LVM and AKS), who rated as case with psychosis or control without psychosis, as well as confidence in rating (confident, not confident). The medical records associated with the admission were manually reviewed for all individuals where there were discordant ratings or rated by at least one rater as not confident. A consensus meeting was held to determine the final classification of discordant/not confident ratings. To compare the rating of psychosis using these phrases with the gold standard of manual review of full medical records, the notes of 470 patients were randomly selected (∼10% of the full sample). The positive predictive value (PPV) of phrase review to classify patients with and without psychosis compared with gold standard medical record review was 97%.

Demographic and clinical characteristics of the overall sample are provided in Table 1, comparing those diagnosed with psychosis (n = 1,196) to controls without psychosis (n = 3,433). Both groups had a similar median age of around 22 years, but the psychosis group featured a higher proportion of males (65.6% versus 47.7%). Racial composition varied, with the psychosis group having a higher representation of Black individuals (11.5% versus 6.9%) and a slightly lower percentage of White individuals (70.7% versus 75.5%). The psychosis group also showed a higher prevalence of public insurance users (17.6% versus 9.2%), indicating possible socio-economic differences. Substance use patterns differed notably between the groups, especially in cannabis and alcohol use, with the psychosis group showing higher daily cannabis use (28.2% versus 14.6%) and lower active alcohol use disorder (21.2% versus 27.5% in controls).

**Table 1.**
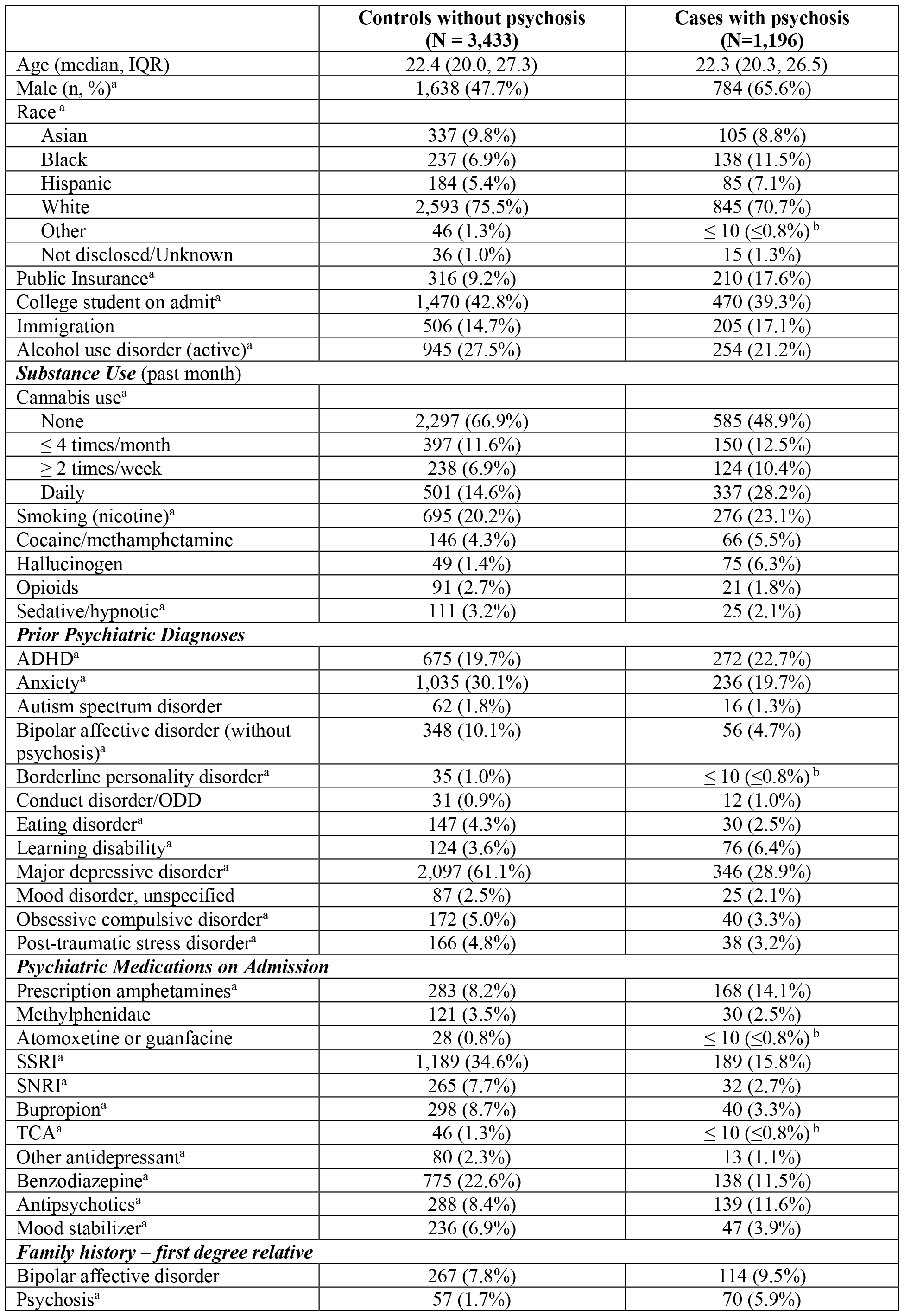
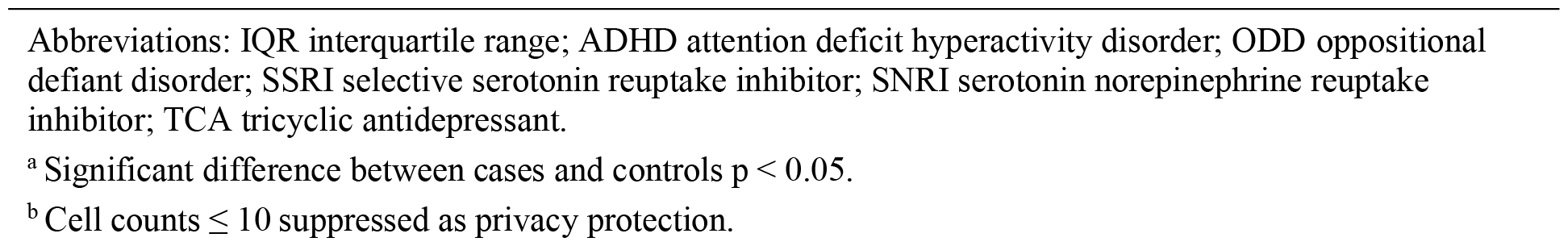
Demographic and Clinical Characteristics of Cases and Controls.

|  | **Controls without psychosis**<br>**(N = 3,433)** | **Cases with psychosis**<br>**(N=1,196)** |
| --- | --- | --- |
| Age (median, IQR) | 22.4 (20.0, 27.3) | 22.3 (20.3, 26.5) |
| Male (n, %)<sup>a</sup> | 1,638 (47.7%) | 784 (65.6%) |
| Race <sup>a</sup> |  |  |
| Asian | 337 (9.8%) | 105 (8.8%) |
| Black | 237 (6.9%) | 138 (11.5%) |
| Hispanic | 184 (5.4%) | 85 (7.1%) |
| White | 2,593 (75.5%) | 845 (70.7%) |
| Other | 46 (1.3%) | ≤ 10 (≤0.8%)<sup>b</sup> |
| Not disclosed/Unknown | 36 (1.0%) | 15 (1.3%) |
| Public Insurance<sup>a</sup> | 316 (9.2%) | 210 (17.6%) |
| College student on admit<sup>a</sup> | 1,470 (42.8%) | 470 (39.3%) |
| Immigration | 506 (14.7%) | 205 (17.1%) |
| Alcohol use disorder (active)<sup>a</sup> | 945 (27.5%) | 254 (21.2%) |
| **Substance Use (past month)** |  |  |
| Cannabis use<sup>a</sup> |  |  |
| None | 2,297 (66.9%) | 585 (48.9%) |
| ≤ 4 times/month | 397 (11.6%) | 150 (12.5%) |
| ≥ 2 times/week | 238 (6.9%) | 124 (10.4%) |
| Daily | 501 (14.6%) | 337 (28.2%) |
| Smoking (nicotine)<sup>a</sup> | 695 (20.2%) | 276 (23.1%) |
| Cocaine/methamphetamine | 146 (4.3%) | 66 (5.5%) |
| Hallucinogen | 49 (1.4%) | 75 (6.3%) |
| Opioids | 91 (2.7%) | 21 (1.8%) |
| Sedative/hypnotic<sup>a</sup> | 111 (3.2%) | 25 (2.1%) |
| **Prior Psychiatric Diagnoses** |  |  |
| ADHD<sup>a</sup> | 675 (19.7%) | 272 (22.7%) |
| Anxiety<sup>a</sup> | 1,035 (30.1%) | 236 (19.7%) |
| Autism spectrum disorder | 62 (1.8%) | 16 (1.3%) |
| Bipolar affective disorder (without psychosis)<sup>a</sup> | 348 (10.1%) | 56 (4.7%) |
| Borderline personality disorder<sup>a</sup> | 35 (1.0%) | ≤ 10 (≤0.8%)<sup>b</sup> |
| Conduct disorder/ODD | 31 (0.9%) | 12 (1.0%) |
| Eating disorder<sup>a</sup> | 147 (4.3%) | 30 (2.5%) |
| Learning disability<sup>a</sup> | 124 (3.6%) | 76 (6.4%) |
| Major depressive disorder<sup>a</sup> | 2,097 (61.1%) | 346 (28.9%) |
| Mood disorder, unspecified | 87 (2.5%) | 25 (2.1%) |
| Obsessive compulsive disorder<sup>a</sup> | 172 (5.0%) | 40 (3.3%) |
| Post-traumatic stress disorder<sup>a</sup> | 166 (4.8%) | 38 (3.2%) |
| **Psychiatric Medications on Admission** |  |  |
| Prescription amphetamines<sup>a</sup> | 283 (8.2%) | 168 (14.1%) |
| Methylphenidate | 121 (3.5%) | 30 (2.5%) |
| Atomoxetine or guanfacine | 28 (0.8%) | ≤ 10 (≤0.8%)<sup>b</sup> |
| SSRI<sup>a</sup> | 1,189 (34.6%) | 189 (15.8%) |
| SNRI<sup>a</sup> | 265 (7.7%) | 32 (2.7%) |
| Bupropion<sup>a</sup> | 298 (8.7%) | 40 (3.3%) |
| TCA<sup>a</sup> | 46 (1.3%) | ≤ 10 (≤0.8%)<sup>b</sup> |
| Other antidepressant<sup>a</sup> | 80 (2.3%) | 13 (1.1%) |
| Benzodiazepine<sup>a</sup> | 775 (22.6%) | 138 (11.5%) |
| Antipsychotics<sup>a</sup> | 288 (8.4%) | 139 (11.6%) |
| Mood stabilizer<sup>a</sup> | 236 (6.9%) | 47 (3.9%) |
| **Family history – first degree relative** |  |  |
| Bipolar affective disorder | 267 (7.8%) | 114 (9.5%) |
| Psychosis<sup>a</sup> | 57 (1.7%) | 70 (5.9%) |

Abbreviations: IQR interquartile range; ADHD attention deficit hyperactivity disorder; ODD oppositional defiant disorder; SSRI selective serotonin reuptake inhibitor; SNRI serotonin norepinephrine reuptake inhibitor; TCA tricyclic antidepressant.
<sup>a</sup> Significant difference between cases and controls $p < 0.05$ .
<sup>b</sup> Cell counts $\leq 10$ suppressed as privacy protection.

## 3. METHODS

Figure 2 illustrates the study design and the intended application of this study. The goal is to assess and identify the most effective NLP model for detecting episodes of psychosis from psychiatric admission notes. Our approach is systematic, starting with the extraction of relevant information from the admission notes using pre-identified keywords. These keywords, rooted in prior research^16^ and refined by our expert consensus, served to sieve through the notes and retain content that was most indicative of psychosis.

**Figure 2.**
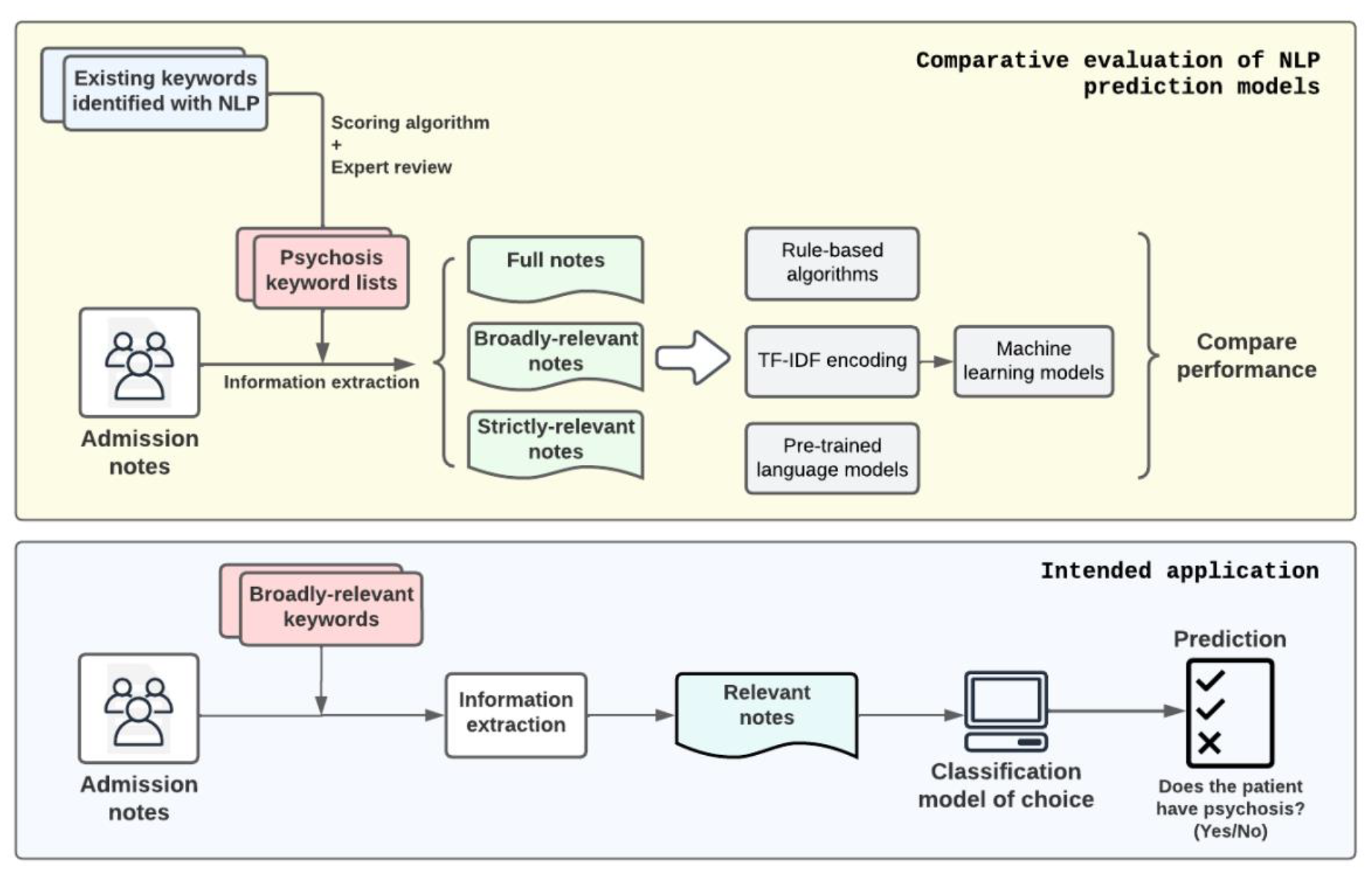
Study Design and Intended Application. The objective of this study is to evaluate and compare the effectiveness of rules-based algorithms, machine learning models, and pre-trained language models in detecting episodes of psychosis from psychiatric admission notes of patients. To address the input length limitations of pre-trained language models and minimize extraneous information, the study incorporates a note pre-selection process. This process involves the use of various keyword sets derived from existing literature, which have been reclassified and reviewed by experts for relevance and accuracy.

Upon preparing the data, we conducted a comparative analysis of three mainstream NLP approaches. The first was a series of rule-based algorithms, which relied on a predetermined set of rules for identifying psychosis. The second approach entailed various machine learning models, which were trained on features extracted by TF-IDF to recognize patterns that might signify psychosis. Lastly, we evaluated pre-trained language models, leveraging their extensive prior training on diverse language datasets, which we fine-tuned for our specific task.

### 3.1. Rule-Based Algorithms

Rule-based NLP algorithms are designed around a set of predefined linguistic rules. These algorithms rely on specific patterns, keywords, phrases, or a combination of linguistic markers that have been historically associated with psychosis. Examples of rule-based approaches that could be employed on our task include keyword identification, pattern matching, semantic rules, etc.

Keyword identification is the simplest NLP method that involves scanning the text for specific keywords or phrases that are strongly associated with psychotic symptoms, such as “hallucinations,” “delusions,” “disorganized speech,” or “catatonia.” The presence of these keywords might trigger a flag for potential psychosis. Pattern matching typically uses regular expressions to find patterns that suggest psychosis. It might look for complex patterns of speech that are indicative of disorganized thinking, a symptom of psychosis, like sentences that lack logical flow or abrupt topic changes. Semantic Rules involves creating rules that not only look for specific words but also their semantic relations. For example, rules that link terms like “hears” with “voices” or “believes” with “being followed” could indicate hallucinations or paranoid delusions, respectively.

In this study, we have chosen to focus on keyword identification, acknowledging that rule-based approaches typically yield suboptimal performance in terms of generalizability. Our objective is to devise a method that maximizes generalization across studies. While both pattern matching and semantic rules could potentially enhance the model’s precision, they necessitate an extensive review of the study corpus to develop bespoke rules. Such customization, however, tends to limit the applicability of the method to other studies and corpora, as the rules may not be universally applicable or effective in different contexts. Therefore, we prioritize broader applicability over corpus-specific optimization in our methodological approach.

We adopted the keyword lists developed in a 2019 study by Viani, et al,^8^ which trained various models on use-case specific EHR texts from early psychosis intervention services, institution-specific discharge summaries, and external clinical texts and also experimented with pre-trained embeddings from MEDLINE/PubMed. Their methodology encompassed diverse data sources, ensuring broad applicability and robustness in generating relevant terms, with the goal of developing an automated NLP model that could be applied to diverse settings outside of McLean Hospital.

The paper presented three keyword sets: the foundational 26 base seed terms related to psychosis symptoms and two generated lists segmented into unigrams and bigrams. Each term from the generated lists was manually categorized as a Relevant Term (RT), a Potentially Relevant Term (PT), or Not Relevant (NT). Guided by this classification, we formulated two distinct rules, which we refer to as “strict” and “broad”, to construct our own keyword lists from the amalgamated unigram and bigram terms:

‐ Strict Rule: A term was considered strictly relevant only if it was unanimously identified as an RT by both annotators.
‐ Broad Rule: A term was deemed broadly relevant if either (1) one annotator marked the term as an RT while the other marked it as a PT or NT, or (2) both annotators agreed on marking the term as a PT.

Following this, we integrated the terms filtered via these criteria with the original seed terms. During post-processing, underscores were substituted with spaces, and plural forms were streamlined. To further reduce computational complexity, both lists were refined to exclusively feature terms with the shortest unique substrings. We also made some minor changes in classification of terms from Viani et al., as annotators, who have clinical experience working with these notes at McLean Hospital, classified notes in accordance with their typical experience (e.g., moved the term “persecutory” from broad to strict because exclusively used in context of describing delusions).

**Table 2.** Psychosis-related keyword lists generated from previous research.

| Base keywords | Strict keywords (excluding base) | Broad keywords (excluding base and strict) |
| --- | --- | --- |
| circumstantial, clang association, delusion, derailment, flight of idea, formal thought disorder, hallucination, loosening of association, paranoia, persecutory idea, psychosi, psychotic, running commentary, somatic passivity, tangential, thought alienation, thought block, thought disorder, thought interference | abnormal belief, abnormal perception, deluded, hallicinat, hallucat, halluciant, halluciat, hallucinat, hallucnat, halluicnat, halucinat, passivity, persecutory, though broadcast, thought blocked, thought broadcast, thought disordered, thought echo, thought insertion, thought withdrawal | aggitation, altered perception, auditory disturbance, bizarre behaviour, bizarre belief, bizarre idea, clanging, delusion-like, delusionary disjoint, disordered, disorganis, echolalia, elated mood, elation, flat affect, ftd, grandiose, guarded, halluc, highly agitated, highly aroused, highly distressed, hostile, hostility, hypomanic, illogical, illusion, incoherence, incoherent, jealousy, knights move, loose association, manic, mute, neologism, nihilistic idea, odd belief, olfactory, over inclusive, over valued, overvalued, paranoid, parnoid, perceptual abnormalit, perceptual disturbance, pressured, referential, religiou, religious theme, running commentarie, seeing shadow, seeing shape, somatic, special abilitie, suspicion, suspicious guarded, suspiciousne, suspiciousne, tactile, tangencial, tangenital, third person, unusual belief, unusual experience, word salad |

### 3.2. Machine Learning Algorithms

Machine learning algorithms are excellent at identifying patterns, but they require data to be in a format that they can process—essentially, numerical. Since textual information is inherently non-numeric, we must convert words into some form of numerical representation, or embeddings, that encapsulate the significance of the words within the context of the document.

#### 3.2.1. Textual inputs to embeddings

There are several methods for encoding textual information, including one-hot encoding, word embeddings like Word2Vec, and the bag-of-words model. Each of these has its strengths, but they might not be the best fit for clinical informatics due to various limitations, such as ignoring word order (bag-of-words) or being computationally intensive (Word2Vec).

We chose the TF-IDF encoding because it provides a balance that is particularly advantageous in the psychiatric domain. TF-IDF quantifies the importance of a word in a collection of documents. It increases with the number of times a word appears in a document (Term Frequency) but is offset by the frequency of the word across all documents (Document Frequency). This means that common words across all documents are deemed less important, while unique words to a document are given more weight.

In our study, we took additional steps to refine the text data for our machine learning models. We removed ‘stopwords’, which are common words like “the”, “is”, and “they” that offer little diagnostic value. We also limited the features to words that appear with a certain frequency—neither too common to be trivial nor too rare to be irrelevant. This was to ensure the words we used as features were statistically significant and had the potential to contribute meaningfully to the diagnosis of psychosis. Our feature set included unigrams (single words), bigrams (pairs of words), and trigrams (three-word phrases) to capture not just the significance of individual words but also the context provided by their adjacent terms. This is important because, in psychiatry, the context in which a word appears can be as telling as the word itself. More details of this process are provided in Appendix A.

#### 3.2.2. Machine learning classification models

Upon transforming psychiatric admission notes into machine-readable embeddings, we used them to develop and evaluate a suite of machine learning classifiers. We included a diverse set of machine learning methods to ensure a comprehensive analysis. This diversity acknowledges that each model’s unique strengths, assumptions, and potential biases play a significant role in its performance in detecting psychosis from textual data. By employing a range of models, we aim to ensure that our findings are robust and not merely an artifact of a single algorithm’s particular tendencies. In detail, we chose four classifiers that are renowned for their efficacy in clinical informatics:

1. **Logistic Regression**. Known for its simplicity and interpretability, estimating the probability of a binary outcome from input features, it operates under the assumption of a linear relationship between the features and the log odds of the outcome and presumes independence between features. However, this can be a potential bias if significant non-linear interactions exist within the psychiatric data, which logistic regression may fail to capture.
2. **Random Forest**. As an ensemble of decision trees that can manage a large number of features, random forest classifiers are adept at classifying complex datasets. However, while they are less prone to overfitting compared to individual decision trees and are good at capturing non-linear relationships, they can still be biased towards more frequent categories or features with more levels, which could overshadow the subtle patterns of psychosis in an unbalanced dataset.
3. **Multilayer Perceptron (MLP)**. As a type of neural network, it is valued for its capacity to learn complex functions through multiple layers of neurons. It assumes that intricate patterns can be discerned through these layers, which is useful for identifying nuanced language patterns in psychiatric notes indicative of psychosis. Nevertheless, biases can arise if the training data isn’t comprehensive, potentially causing the MLP to overlook less common but clinically relevant expressions of psychosis.
4. **XGBoost**. Leveraging a boosting algorithm that builds models sequentially to correct prior errors, XGBoost assumes that continuous learning from mistakes enhances performance. While known for its accuracy and efficiency, it can become biased by overemphasizing outliers or noisy data, and without careful tuning, it may overfit, learning the training data too well and failing to generalize to new, unseen data.

### 3.3. Pre-trained Language Models

Pre-trained language models are a significant advancement in the field of deep learning, which is a subset of machine learning focused on algorithms inspired by the structure and function of the brain called artificial neural networks. The term “pre-trained” refers to the process where these models have already learned a substantial amount of English language understanding before they are fine-tuned for a specific task—much like a medical student who has gone through years of training before specializing. The pre-training equips these models with a deep knowledge of language structure and word relationships, allowing them to generate embeddings—numeric representations of text that capture semantic meaning.

In practice, these models serve primarily as sophisticated embedding generators that translate textual data into a numerical form, rather than acting directly as classifiers. The rich embeddings they create are versatile and can be utilized in a range of downstream tasks, prediction being just one example. Given the complexity and depth of understanding inherent in these pre-trained models, the classifiers used in conjunction with them can be relatively straightforward. A common choice is a linear classification layer, which, despite its simplicity, is sufficient for making decisions based on the comprehensive information contained within the embeddings.

In this study, we chose Bidirectional Encoder Representations from Transformers (BERT)^17^ models, which strike a balance between size, computational efficiency, and performance. While there are larger language models available, known for their robust performance in processing human languages, BERT’s architecture offers a more practical alternative for clinical research environments. The latest large language models, though powerful, require considerable computational resources for fine-tuning and inference processes—resources that many clinical labs may not have. BERT models, being medium-sized, demand less in terms of computing power and data for training, which aligns better with the typical resource constraints found in clinical settings.

We utilized specialized BERT variants, namely ClinicalBERT^18^ and BlueBERT,^19^ which have been pre-trained on extensive medical corpora, including MIMIC-III and PubMed. This pre-training imbues the models with an inherent understanding of medical terminology and documentation structure, allowing them to excel at identifying clinical conditions from text data, such as signs of psychosis in patient notes. Their pre-existing familiarity with medical lexicon and semantic constructs positions them as efficient tools for parsing clinical notes, providing a significant advantage over models that have not been specialized, such as generic language models or those trained on non-medical text.

For both models, we implemented the architectures using all BERT embedding layers while freezing all but the last layer. To tailor the models for the task of psychosis identification, we added a linear classification layer. Fine-tuning was conducted with all but the last embedding layer and the added linear classification layer fixed to avoid overfitting.

Pre-trained language models like BERT use a self-attention mechanism which computes the attention scores for each pair of tokens (i.e., chunks of words) in the input sequence and thus requires computational resources that grow quadratically with the increase in input length. The two BERT models we use both set the maximum number of tokens per input. Therefore, for inputs with lengths more than 512 tokens, we truncate them to keep the first 512 tokens. More experiment settings regarding these models can be found in Appendix C.

### 3.4. Experiment design

#### 3.4.1. Training and evaluation details

As depicted in Figure 3, the lengths of psychiatric admission notes are both long and variable, often surpassing the token limitation imposed by BERT models, and potentially introducing noise into machine learning models. Consequently, we applied the three-tiered keyword lists developed during the rule-based method phase as an initial filtering step before inputting data.

**Figure 3.**
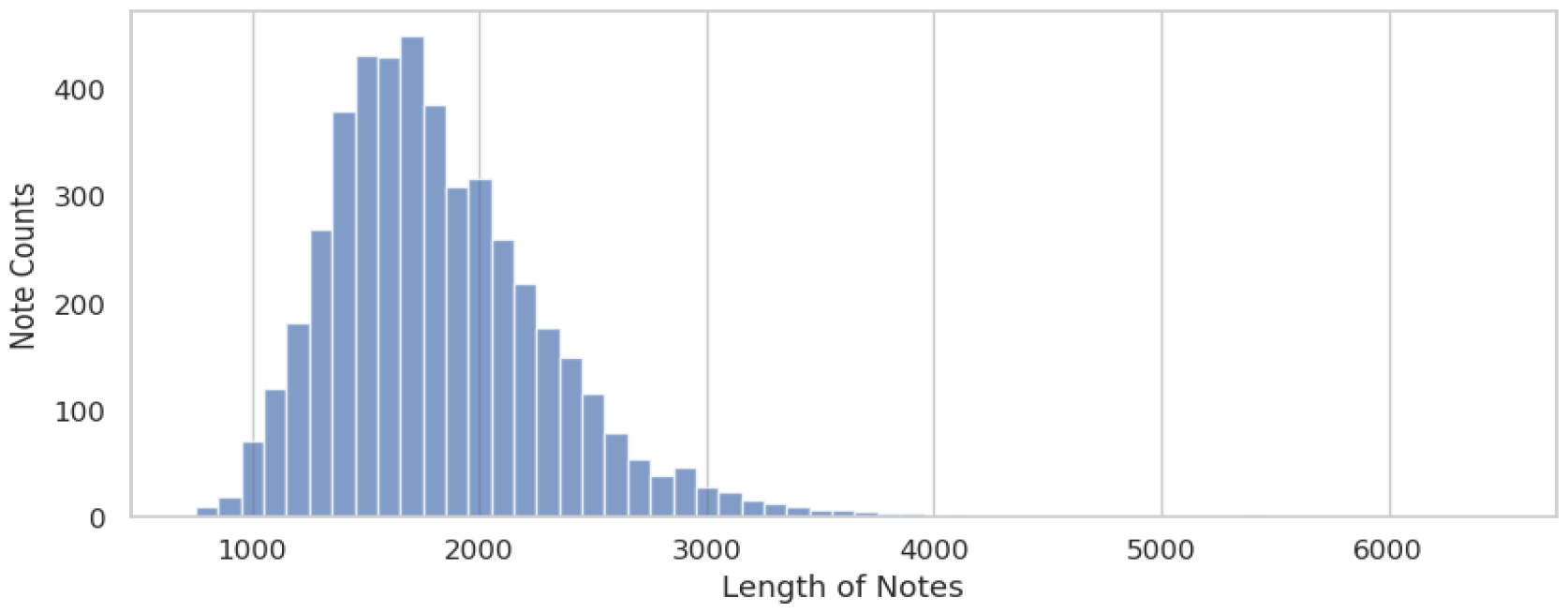
Length distribution of admission notes in our dataset, split by whitespaces.

Prior to the experiments, the dataset was partitioned into training, validation, and test subsets. The training set’s purpose is to train both machine learning and pre-trained language models, while the validation set is utilized to test the models during the training phase. This intermediate subset is crucial as it enables the model to iteratively make predictions and monitor the progression of the learning process. The test subset acts as a final, consistent measure for validating the performance of all models.

We prepared three distinct versions of the dataset using the keyword lists: (1) the full notes, (2) notes compiled from sentences containing broadly relevant keywords, and (3) notes assembled from sentences with base relevant keywords. We omitted creating a set for strictly relevant keywords because every psychiatric admission note contained at least one base keyword. Thus, our comparison was between the base keywords (the most pertinent as determined by psychiatrists) and the broadly relevant keywords (expert-reviewed keywords identified by NLP methods).

For rule-based algorithms, which do not require training, evaluation was conducted directly on the test set. Machine learning algorithms, not constrained by input length, were assessed using the full notes as well as the sets filtered by broadly relevant and base keywords. Pre-trained language models, due to their limited input length capacity, were only applied to the sets of broadly relevant and base keywords, as truncating full notes to the first 512 tokens could result in the loss of critical information.

#### 3.4.2. Evaluation methods and metrics

We used the F1 score, the harmonic means of precision (positive predictive value [PPV]) and recall (sensitivity), as the primary metrics for evaluating our models’ performance. We also reported the mean and standard deviation of sensitivity, specificity, PPV, negative predictive value (NPV), and accuracy. This allowed us to assess the stability and robustness of our results and determine the overall performance of the algorithms.

For the machine learning algorithms and pre-trained language models, we used a bootstrap resampling strategy with 1000 samples to estimate the confidence interval of the performance metrics. In addition, we report their area under the receiver operating characteristic curve (AUROC), area under the precision-recall curve (AUPRC), and respective 95% confidence intervals (CI).

#### 3.4.3. Non-NLP baseline: ICD code identification

We used the International Classification of Diseases (ICD) codes from discharge notes, which represented the principal diagnosis at discharge, as our baseline to compare NLP-based methods. Notably, discharge notes are expected to offer higher diagnostic accuracy compared to admission notes because they encompass the entirety of a patient’s hospital stay, including all diagnoses, treatments, diagnostic clarification, and outcomes. The rule-based algorithm is straightforward but highly specific, employing an exact-match approach. When a patient’s discharge notes include any ICD-9 or ICD-10 codes encompassing psychosis, the algorithm flags that patient as having psychosis. Conversely, if these codes are absent from the discharge notes, the patient is not considered to have psychosis.

The ICD codes considered are comprehensive, covering a wide array of psychosis-related conditions. These include, but are not limited to, *Psychosis, unspecified* (F28, F29); *Non-organic, other transient psychoses* (F23); *Schizophrenia spectrum disorders* (F20, F25); and *Substance-induced psychoses such as Cannabis-induced psychosis* (F12.15, F12.25, etc.) and *Stimulant-induced psychosis* (F15.15, F15.25, etc.). The exhaustive list can be found in Appendix B.

## 4. RESULTS

Table 3 shows the comparative evaluation results. The most effective methods per metric are highlighted in green, and the least effective in red. The F1 score was employed as the primary metric to ascertain the optimal model or algorithm.

**Table 3.**
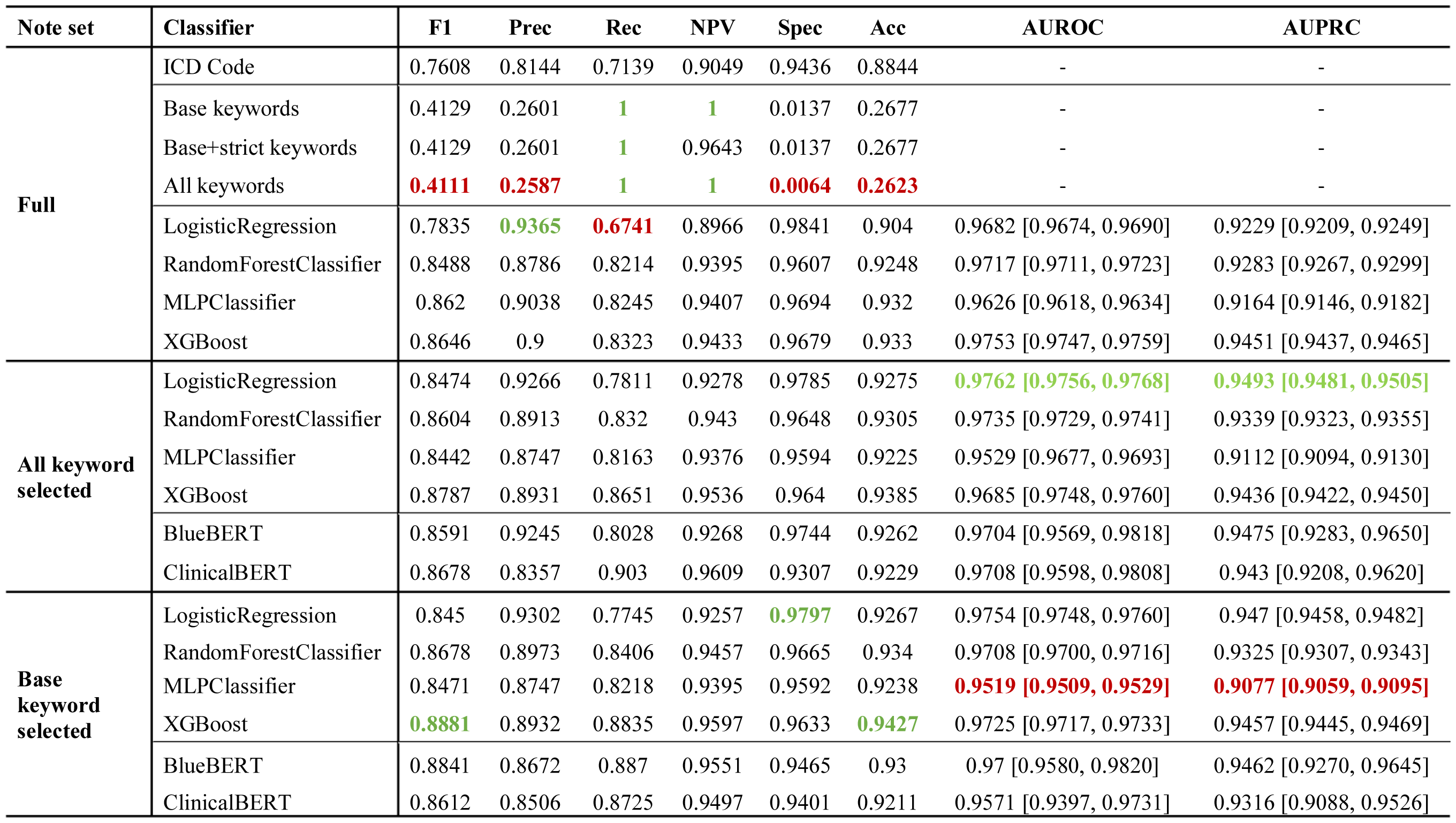
Comparative Analysis of Algorithm Performance in Psychosis Detection from Psychiatric Admission Notes. “Full note” represents notes subjected only to the preprocessing stage. “All keyword-selected” refers to notes compiled from sentences marked by any keyword from the base, strict, or broad keyword lists; “Base keyword-identified” pertains to notes composed of sentences flagged by keywords in the base list. The most effective methods per metric are highlighted in green, and the least effective in red. The F1 score was employed as the primary metric to ascertain the optimal model or algorithm.

We observe that the non-NLP-baseline ICD identification exhibited reasonable performance, with an F1 score of 0.761, a recall of 0.714, and an NPV of 0.905. While its specificity and accuracy were relatively high at 0.944 and 0.884, respectively, it lagged in precision at 0.814.

The keyword-matching algorithms, particularly those based on all keyword lists, demonstrated exceptionally high recall and NPV scores, both reaching the maximum value of 1.0 at the cost of extremely low precision (around 0.26) and specificity (as low as 0.0064). Nonetheless, the keywords served as great pre-selection methods, as we observe that is that most of the models had performance gains going from full notes to base-keyword identified notes. Appendix D shows summary statistics of keyword distribution in notes of cases of psychosis and controls.

The machine learning methods, TF-IDF + XGBoost demonstrated the best performance, achieving F1 scores of 0.8646, 0.8787, and 0.8881 on the three sets of notes, respectively. The model also recorded the highest accuracy of 0.9330, 0.9385, and 0.9427 on the full and selected sets. This consistent outperformance of other models underscores XGBoost’s robustness across different feature spaces and its ability to balance precision and recall effectively.

One noteworthy observation relates to the performance of the logistic regression model, particularly in terms of precision. On the full dataset, the logistic regression model achieved a precision of 0.9365 and a specificity of 0.9841, the highest among all classifiers in the category using full notes. Nonetheless, logistic regression models were low in recall (sensitivity) scores (0.6741).

Both BlueBERT and ClinicalBERT demonstrated high AUROC and AUPRC, reflecting their capabilities in exceptional class differentiation and classification. However, their AUROC and AUPRC are subject to greater fluctuation in 95% confidence intervals compared to XGBoost.

As of our key metric of F1 score, BlueBERT trained and tested on admission notes pre-selected using only the base keywords achieves a remarkable F1 score of 0.8841. This surpasses the version trained and tested on admission notes pre-selected using all available keywords, which posts an F1 score of 0.8591. BlueBERT trained on base keyword identified notes demonstrates stronger precision and specificity, reinforcing the idea that careful keyword selection helps reduce noise and enhance model performance.

## 5. DISCUSSION

Our study comprehensively evaluated NLP methods in identifying psychosis in psychiatric patients through a multi-pronged analysis involving rule-based algorithms, machine learning, and pre-trained language models using psychiatric admission notes from 4,629 patients. We found that keyword pre-selection of notes increased classifier performance, XGBoost using TF-IDF-encodings performed the best among all methods, BlueBERT offered similar performance. A previous study^12^ that predicted conversion to psychosis among patients with a psychiatric disorder identified 14 terms that predicted psychosis, which included non-specific symptoms of psychiatric illness (e.g., insomnia), substance use (e.g., cannabis and cocaine use), and symptoms more specific to psychosis (e.g., delusions). Symptoms more relevant to psychosis, specifically delusions, agitation, and paranoia, were the strongest predictors of psychosis. This study used supervised machine learning (support vector machine) based on a lexicon of keywords indicating symptoms of serious mental illness. Our study adds to this research by showing that deep learning methods can enhance the performance of models to identify psychosis among patients admitted to a psychiatric hospital from electronic health records.

In our evaluation, keyword-based algorithms, especially those using a less curated set of keywords, demonstrated extremely unbalanced performance metrics. These algorithms achieved essentially perfect recall but had very poor precision (i.e., positive predictive value), as low as 0.2587. Such results indicate that while the keyword-based algorithms are adept at identifying relevant cases, they also produce a high number of false positives. A significant drawback of such algorithms lies in their inability to grasp context, particularly when it comes to negations and expressions of uncertainty; phrases like “no signs of psychosis” can trigger false positive results. This deficiency in contextual comprehension underscores the necessity for more sophisticated NLP techniques or machine learning models capable of grasping the semantics of clinical language.

Nonetheless, the base keywords played a pivotal role in pre-selecting relevant information from clinical admission notes, thereby streamlining the performance of both machine learning classifiers and pre-trained language models. By filtering out extraneous details, the keyword-based selection process not only made the classifiers more efficient but also optimized pre-trained language models to work within input size constraints. This dual utility enhanced diagnostic accuracy and computational efficiency across the board. Interestingly, however, we discovered that incorporating additional relevant keywords extracted from extensive PubMed texts and clinical notes using word embedding techniques did not lead to further improvements in the predictive models’ performance. This discovery may be attributable to the fact that, with the foundational keyword lists that include more directly related keywords, the matched text already reached the input limit. As our cohort was identified in a psychiatric hospital with high rates of psychosis, applying our model to the general population may be more challenging due to rarity of symptoms of psychosis. However, as the keyword lists adopted from Viani et al.^16^ used diverse data sources, this comprehensive list of keywords may have applicability in other settings. In addition, many of the signs and symptoms of psychosis are domain-specific, such that relevant terms were curated by domain experts using their clinical experience.

Among the machine learning classifiers we tested, XGBoost emerged as the most promising, achieving an F1 score of 0.8881. Remarkably, the best-performing pre-trained language model exceeded all other models, boasting an F1 score of 0.8841. This represents an increase from the ICD codes’ F1 score of 0.7608, which is noteworthy since ICD codes reflected diagnosis at discharge and may be more accurate as they incorporate information gained from observation and clinical evaluation over the course of days to weeks.

The use of keyword-based selection in tandem with machine learning and pre-trained language models offers a particularly promising avenue for identifying specific patient cohorts in neuropsychiatric epidemiological studies. Traditional methods of cohort identification often rely on manual curation and are thus time-consuming and subject to human error. The method used for cohort identification for this study by clinicians, which included review of phrases related to psychosis and manual review of a subset of medical records, was labor-intensive and would not be feasible for larger samples. The NLP techniques presented in this paper could automate and significantly expedite this process, allowing researchers to rapidly and accurately isolate groups of patients who share certain diagnostic features, treatment histories, or risk factors. This level of granularity is crucial for epidemiological studies that aim to understand the complex interplay of various factors in neuropsychiatric conditions. For example, researchers could more easily identify cohorts for longitudinal studies on treatment efficacy or for cross-sectional studies aimed at identifying environmental or genetic risk factors. Overall, the technological advancements discussed in the paper could revolutionize the way patient cohorts are identified and analyzed, thereby increasing the speed and accuracy of neuropsychiatric research. Automation of detection of psychosis in EHR notes could be useful clinically to identify patients with emerging psychosis, which currently relies on patient-initiated help-seeking^8^ and specific referral pathways for people at risk for suspected psychosis.^9^ Studies have shown that these current detection strategies are highly inefficient and unreliable, with only 5-12% of individuals at clinical high risk for psychosis (CHR-P) actually converting to first-episode psychosis.^20^ Our methods could allow for detection of at-risk individuals across a broad hospital system, allowing triage to more intensive level of care or refer to clinics that provide comprehensive services for individuals with first-episode psychosis.

However, the use of admission notes as the primary data source also has limitations. These notes are subject to clinician bias and incomplete information about patients. For example, patients may be too agitated, sedated from emergent medications or lack insight into symptoms to provide accurate information. Algorithmic bias may be introduced by clinician bias, as previous studies have shown that Black individuals with depression are more likely to be misdiagnosed with psychosis than White patients.^21^ Additionally, the absence of objective, quantifiable biomarkers for mental health conditions further complicates diagnostic accuracy. Signs of mental health conditions may not be immediately evident and may only emerge over time, affecting the classifiers’ performance, particularly in terms of specificity and precision. Therefore, these models should be part of a broader diagnostic toolkit, possibly incorporating longitudinal data, in-depth interviews, and other objective measures for a more comprehensive understanding of a patient’s mental health.

Ethical considerations also arise, particularly regarding the potential for overdiagnosis or misdiagnosis, given the limitations of both keyword-based and machine-learning algorithms. Such risks necessitate rigorous validation and possibly the inclusion of human oversight in the diagnostic process.

In conclusion, while machine learning algorithms offer promise for improving mental health diagnosis, further research is required to address their limitations and to explore their integration into a more comprehensive diagnostic framework. We recommend that future studies consider incorporating more diverse and longitudinal data sources to validate and potentially improve upon our findings.

## 6. CONCLUSION

This study presents a comprehensive evaluation of common NLP techniques for identifying psychosis in from psychiatric admission notes, highlighting the potential of keyword pre-selection and advanced algorithms to refine diagnosis from EHR data. It serves as a guidebook for future studies in using NLP to identify psychosis from EHR. This study also underscores the need for further research to optimize these NLP approaches, aiming for their integration into a holistic diagnostic framework that can augment the capabilities of mental health professionals.

## ACKNOWLEDGEMENTS

This work was supported by NIMH grant R01MH122427.

## AUTHOR CONTRIBUTION

Credit authorship contribution statement:

Conceptualization: Yining Hua, Lauren V. Moran, Li Zhou,

Data Curation, Annotation: Yining Hua, Ann K. Shinn, Lauren V. Moran, Joseph P. Skinner

Methodology and Implementation: Yining Hua

Formal analysis: Yining Hua

Validation: Lauren V. Moran, Joseph P. Skinner

Writing - Original Draft: Yining Hua and Suzanne V. Blackley

Writing – Revision Draft: All authors

Yining Hua takes the responsibility for the integrity of the work.

## DATA AVAILABILITY

We received a data sharing exemption for the NIH grant funding this study, as the Mass General Brigham Institutional Review Board deemed that individual subject-level data could not be shared as patients did not provide informed consent in accordance with waiver of consent for study, as well as increased protection of information on vulnerable populations, including patients with psychiatric disorders.

## APPENDICES Appendix A. TF-IDF embedding settings

Given the imbalanced nature of our dataset—approximately a 1:3 ratio of cases to controls— we used the following steps for refining the feature space. We first excluded standard English stopwords (e.g., high frequency words and words with low information value, such as “the”, “is” and “they”) provided by the Gensim library. To sharpen the model’s attention on distinctive words that hold more diagnostic promise for distinguishing between different documents, we introduced a threshold for feature frequency. We decided that any word found in more than one-third of the documents would be too common to offer any unique insights and thus excluded such words from the TF-IDF matrix. Concurrently, to ensure the statistical significance of our features, we considered only those terms that featured in at least 40 documents, which corresponded to approximately 1% of our sample size. This threshold was pivotal in balancing the need to avoid overfitting with very rare terms and underfitting with overly common terms.

### Appendix B. ICD codes used in identifying patients with psychosis

Appendix B. ICD codes used in identifying patients with psychosis.
| Psychosis code | ICD-9 | ICD-10 |
| --- | --- | --- |
| Psychosis, unspecified | 298.8, 298.9 | F28, F29 |
| Non-organic, other transient psychoses/ brief psychotic disorder | 298.0, 298.1, 298.4 | F23 |
| Psychosis due to known physiological condition | 293.81, 293.82 | F06.0, F06.2 |
| Puerperal/shared psychotic disorder | 297.3 | F24, F53 |
| Hallucinations | 780.1 | R44.0, R44.1, R44.2, R44.3 |
| Delusional disorder | 297.x | F22 |
| Schizophrenia spectrum disorders | 295.x | F20, F25 |
| Alcohol-induced psychosis | 291.3, 291.5 | F10.15, F10.150, F10.151, F10.159, F10.25, F10.250, F10.251, F10.259, F10.95, F10.950, F10.951, F10.959 |
| Opioid-induced psychosis |  | F11.15, F11.150, F11.151, F11.159, F11.25, F11.250, F11.251, F11.259, F11.95, F11.950, F11.951, F11.959 |
| Cannabis-induced psychosis |  | F12.15, F12.150, F12.151, F12.159, F12.25, F12.250, F12.251, F12.259, F12.95, F12.950, F12.951, F12.959 |
| Sedative-hypnotic induced psychosis |  | F13.15, F13.150, F13.151, F13.159, F13.25, F13.250, F13.251, F13.259, F13.95, F13.950, F13.951, F13.959 |
| Cocaine-induced psychosis |  | F14.15, F14.150, F14.151, F14.159, F14.25, F14.250, F14.251, F14.259, F14.95, F14.950, F14.951, F14.959 |
| Other stimulant-induced psychosis |  | F15.15, F15.150, F15.151, F15.159, F15.25, F15.250, F15.251, F15.259, F15.95, F15.950, F15.951, F15.959 |
| Hallucinogen-induced psychosis |  | F16.15, F16.150, F16.151, F16.159, F16.25, F16.250, F16.251, F16.259, F16.95, F16.950, F16.951, F16.959 |
| Inhalant-induced psychotic psychosis |  | F18.15, F18.150, F18.151, F18.159, F18.25, F18.250, F18.251, F18.259, F18.95, F18.950, F18.951, F18.959 |
| Other drug-induced psychosis | 292.1, 292.11, 292.12 | F19.15, F19.150, F19.151, F19.159, F19.25, F19.250, F19.251, F19.259, F19.95, F19.950, F19.951, F19.959 |
| Major depressive disorder with psychotic features | 296.24, 296.34 | F32.3, F33.3 |
| Bipolar disorder, mania/mixed with psychotic features | 296.04, 296.14, 296.44, 296.64 | F30.2, F31.2, F31.64 |
| Bipolar disorder, depressed with psychotic features | 296.54 | F31.5 |

\*ICD9/ICD-10 psychosis codes used for outcome. -x means any digit.

### Appendix C. Experiment details of ClinicalBERT and BlueBERT

For implementation, we employed the Hugging Face API, leveraging the PyTorch^22^ library in Python for the backend computation. To address the skewed distribution of our data, we adapted the loss function to a weighted cross-entropy form, calibrated based on the inverse frequencies of the classes in our dataset. This ensures that the minority class receives a balanced treatment during the training phase. We followed a robust training scheme, which consisted of 25 training epochs, a weight decay of 0.01, and a warm-up phase consisting of 500 steps. Early stopping mechanisms were employed to prevent overfitting, with a patience parameter set to 3. Due to ClinicalBERT having a lighter weight compared to BlueBERT, we set the training and evaluation batch sizes to 32 for ClinicalBERT and 12 for BlueBERT.

### Appendix D. Keyword distribution and note lengths of the admission notes

Table 5 provides insights into the keyword distribution and note lengths of the admission notes shortened by selecting sentences with at least one keyword. Notes associated with psychosis had a median of 12 base keywords, compared to just 5 in notes without psychosis, demonstrating that our keyword list successfully homes in on more clinically relevant details. A similar trend is seen for all keywords, with a median count of 48 for psychosis notes versus 19 for others. The median length of the notes, restricted to sentences that included any keywords, also varies significantly at 374 words for psychosis cases and 154 for non-psychosis cases.

|  | With psychosis | Without psychosis | All |
| --- | --- | --- | --- |
| Median (std) base keywords in each note | 12.0 (6.5) | 5.0 (3.78) | 6.0 (5.72) |
| Median (std) all keywords in each note | 48 (23.86) | 19 (13.73) | 24 (21.26) |
| Median (std) length of each note (all keywords) | 374 (333.57) | 154 (305.7) | 211 (324.63) |
| Median (std) length of each note (base keywords) | 285.5 (289.45) | 88 (253.23) | 153 (273.82) |

